# Asthma Exacerbation Prediction and Interpretation based on Time-sensitive Attentive Neural Network: A Retrospective Cohort Study

**DOI:** 10.1101/19012161

**Authors:** Yang Xiang, Hangyu Ji, Yujia Zhou, Fang Li, Jingcheng Du, Laila Rasmy, Stephen Wu, Wenjin.Jim Zheng, Hua Xu, Degui Zhi, Yaoyun Zhang, Cui Tao

## Abstract

**Background:** Asthma exacerbation is an acute or sub-acute episode of progressive worsening of asthma symptoms and can have significant impacts on patients’ daily life. In 2016, 12.4 million current asthmatics (46.9%) in the U.S. had at least one asthma exacerbation in the previous year.

**Objective:** The objectives of this study were to predict the risk of asthma exacerbations and to explore potential risk factors involved in progressive asthma.

**Methods:** We proposed a time-sensitive attentive neural network to predict asthma exacerbation using clinical variables from electronic health records (EHRs). The clinical variables were collected from the Cerner Health Facts® database between 1992 and 2015 including 31,433 asthmatic adult patients. Interpretations on both the patient level and the cohort level were investigated based on the model parameters.

**Results:** The proposed model obtains an AUC value of 0.7003 through 5-fold cross-validation, which outperforms the baseline methods. The results also demonstrate that the addition of elapsed time embeddings considerably improves the performance on this dataset. Through further analysis, it was witnessed that risk factors behaved distinctly along the timeline and across patients. We also found supporting evidence from peer-reviewed literature for some possible cohort-level risk factors such as respiratory syndromes and esophageal reflux.

**Conclusions:** The proposed time-sensitive attentive neural network is superior to traditional machine learning methods and performs better than state-of-the-art deep learning methods in realizing effective predictive models for the prediction of asthma exacerbation. We believe that the interpretation and visualization of risk factors can help the clinical community to better understand the underlying mechanisms of the disease progression.

## Introduction

Asthma is a common and serious health problem which affects 235 million people worldwide[1] and an estimated of 26.5 million people (8.3% of the U.S. population) in the U.S.[2]. Asthma takes a significant toll on the population which imposes an unacceptable burden on health care systems with a total annual cost of $81.9 billion in 2013 in the U.S. [3,4]. Asthma may develop into exacerbation if it is not well controlled or stimulated by specific risk factors[3]. In 2016, 12.4 million current asthmatics (46.9%) in the U.S. had at least one asthma exacerbation in the previous year[2]. Exacerbations of asthma can be severe and require immediate medical interventions, either as an emergency department visit or an admission to hospital[5]. Serious asthma exacerbations may even result in death[6]. Therefore, it is of practical significance to make early predictions so that interventions can be carried out in advance to reduce the probability of exacerbation.

Investigations on prediction and risk factor recognition for asthma exacerbation have been respectable, in which the main stream adopts traditional statistical methods, such as logistic regression[7,8], proportional-hazards regression[9], and generalized linear mixed models[10]. However, most of them have only explored a small group of candidate risk factors, are usually hard to extend to other datasets and hard to make personalized predictions. With the explosion of healthcare data in recent years, machine learning methods have grown in prominence for this domain, due to their superiority over statistic methods in processing larger numbers of variables and capacity in mining more possible correlations between them[11]. Typical models include Naïve Bayes[13], Bayesian networks[12–14], artificial neural networks[12], Gaussian Process[12], and Support Vector Machines[12,13]. Although different attempts have been made, there are still several deficiencies in applying these traditional machine learning methods. For example, ignoring temporal dependencies between variables might not provide a meaningful risk estimation of future exacerbations for individual patients[15]. Furthermore, most approaches only concentrate on the performance but lack further attention to personalized risk factors.

Recent predictive modeling-related studies focused more on deep learning, which has an upper hand on healthcare predictions because of its flexibility in dealing with longitudinal data[16], powerful learning capabilities[17], and ability to tackle the problem of data irregularity[18]. One of the most popular architectures of deep learning-based predictive model is the recurrent neural networks (RNNs), which take a patient’s visit sequence as the input and make predictions according to the encoded representations. Multiple successes have been achieved in applying deep learning on disease prediction[19], mostly using variants of RNNs with distinct network components, such as attention mechanism for evaluating weights of each variable[20–22], and special configurations in tackling the problem of time decays[20,21,23,24]. Typical prediction tasks include the prediction of diabetes mellitus[18], Parkinson[18,25], chronic heart failure[21], sepsis[26], mortality and readmission[20].

Inspired by previous studies, we applied Long Short-term Memory (LSTM), a popular RNN variant used by dozens of previous studies[19,20,23,27], for asthma exacerbation prediction. We proposed the **T**ime-**S**ensitive **A**ttentive **N**eural **N**etwork (TSANN), which employs a self-attention mechanism[28] to help model the context of both visit-level and code-level variables. Meanwhile, to incorporate the impact of elapsed time, we projected the visit time of each clinical variable into a low-dimensional space and assigned a numeric vector to each time. Making use of the attention weights of TSANN, data analysis was then conducted to investigate personalized and cohort-level risk factors.

As far as we know, this is the first data-driven study to predict asthma exacerbation using deep learning and EHR data, the first effort to introduce elapsed time embeddings into clinical predictive modeling, and the first attempt to visualize risk factors of asthma exacerbation on both the individual level and the cohort level, which have been insufficiently explored in most previous studies. We believe that the outcomes of this study can help the clinical community to better understand the underlying mechanisms of the disease progression and to assist in decision-making. Although focusing on asthma exacerbation for this specific project, the proposed approach can also be adopted in risk prediction for other chronic diseases.

## Methods

### Database

This study used Cerner Health Facts®, a HIPAA-compliant database collected from multiple enrolled clinical facilities, containing mostly in-patient data. Data in Health Facts were extracted directly from the EHRs from hospitals with which Cerner has a data use agreement. Encounters may include the pharmacy, clinical and microbiology laboratory, admission, and billing information from affiliated patient care locations. All personal identifying information of the patients was anonymized. In our study, we primarily focused on the impact of clinical factors on asthma exacerbation, so we extracted diagnosis, medications, and demographic characteristics such as gender, race, and age from the database. The University of Texas Health Science Center (UTHealth) had agreements with Cerner to use this data for research purposes. The institutional review board at UTHealth approved the study protocol.

### Study Design

We conducted a retrospective study to predict the risk of asthma exacerbation. We extracted patients’ records between 1992 and 2015 from the Cerner database. For clarity, we define several terms in advance (Table 1).

**Table 1.**
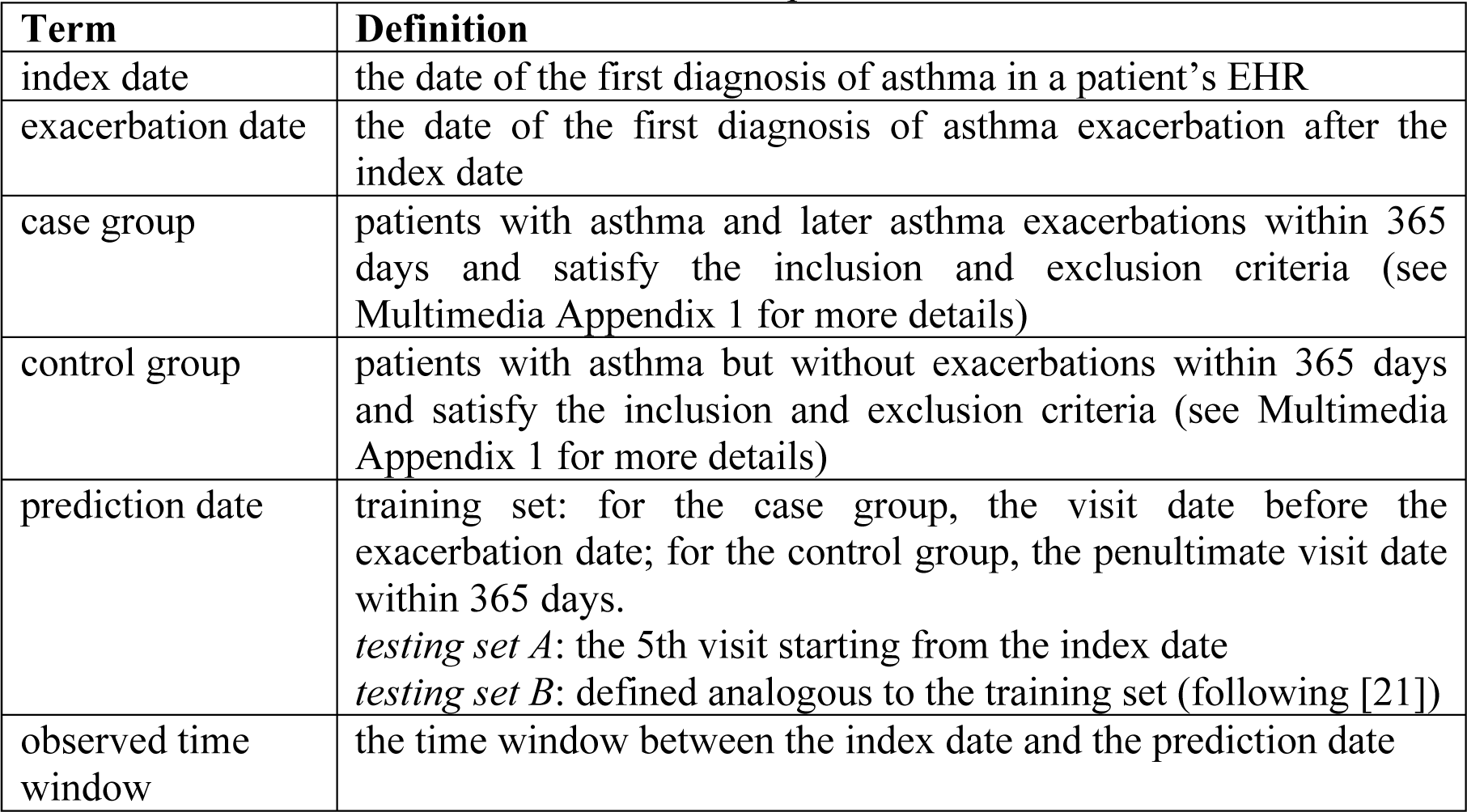
Defined terms for asthma exacerbation prediction.

Intuitively, a testing sample should be defined in a similar way as that for the training samples (as *testing set B*). However, since we cannot foresee when the exacerbation would happen in real-world deployment, we can only make future predictions at each visit. In our study, we selected the 5^th^ visit from asthma index as the prediction date (*testing set A*), considering both leveraging more visits and keeping more patients for experiments (see Multimedia Appendix 1 for more details). We also defined *testing set B*: the penultimate visit as the prediction date, behaving as the upper bound of the classifier performance, since it enables more complete visit information.

The TSANN model was trained to predict the onset of asthma exacerbation given the observed time window. The main outcomes of the method are: (1) a score that measures the risk of asthma exacerbation for each patient; (2) visualization of the results including a personalized heatmap identifying the importance of each clinical variable in the observed time window, cohort-level risk factors and their temporal distributions among patients. Based on the outcomes, further data mining or clinical trials can be carried out for validation. The workflow of this research is shown in Figure 1.

**Figure 1.**
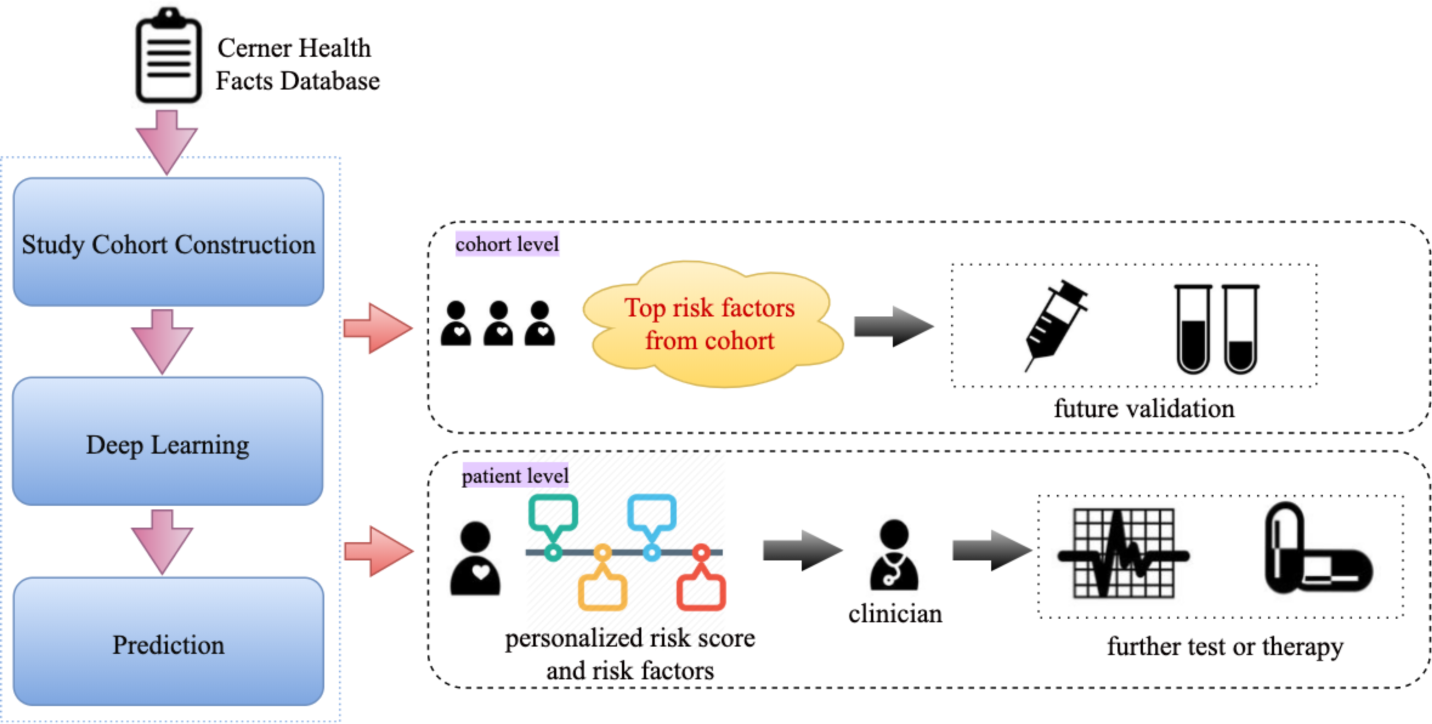
The workflow of risk prediction of asthma exacerbation.

### Selection of Study Subjects

The subjects in the study were patients with a diagnosis of asthma. Inclusion and exclusion criteria were decided based on previous work[29,30] including the diagnosis of asthma and asthma exacerbation. The current study only focused on adult patients with age between 18 and 80. In the end, 31,433 individuals remained, including 2,262 cases and 29,171 controls (≈1:13). The cohort selection process is shown in Figure 2. More details for the cohort selection are shown in the Multimedia Appendix 1.

**Figure 2.**
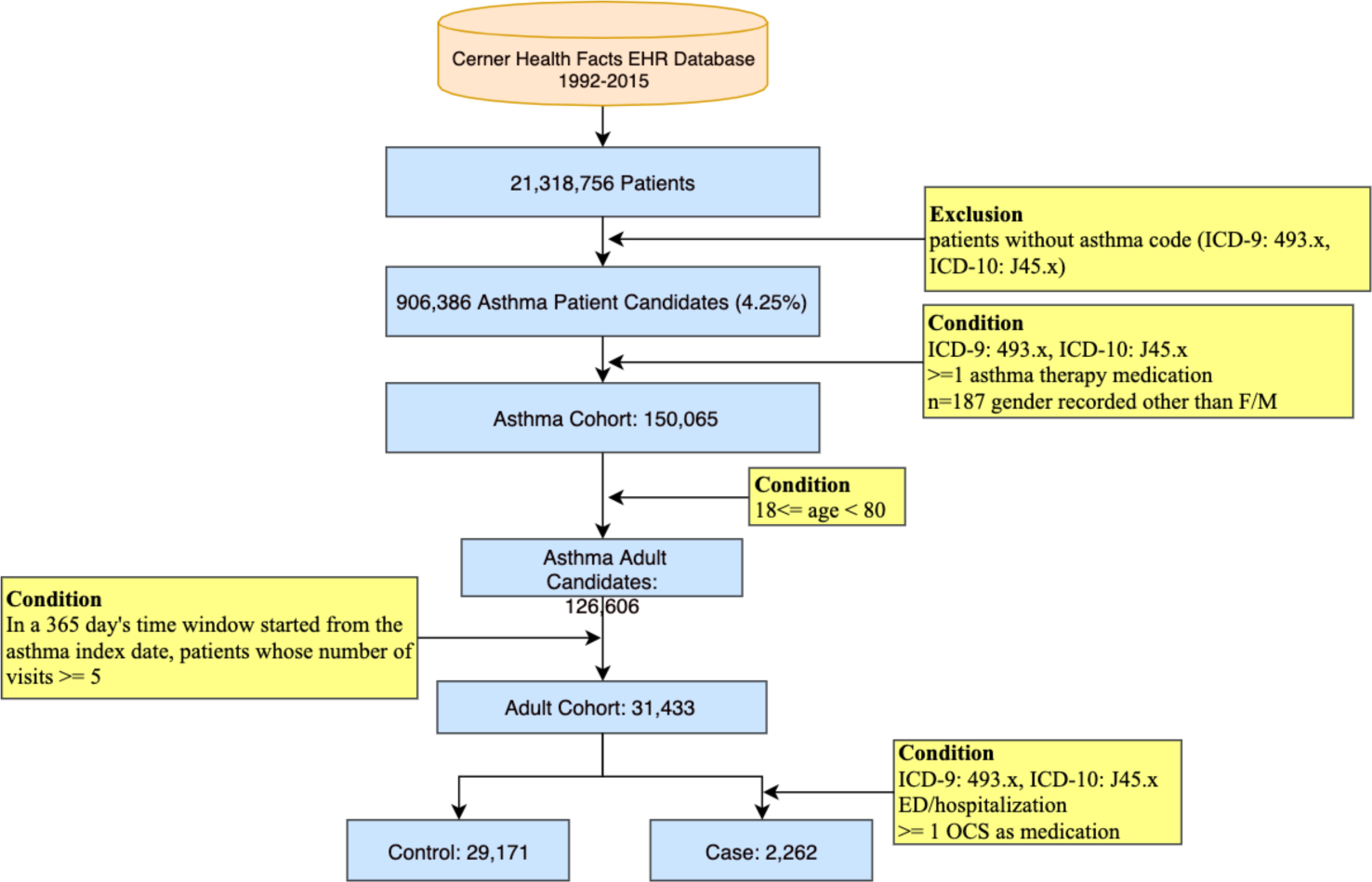
The cohort selection process for the study of asthma exacerbation.

### Time-sensitive Attention Neural Network

#### Model Overview

TSANN accepts the whole sequence of clinical variables in the observed time window as inputs, and outputs the probability of asthma exacerbation (see Figure 3). The architecture of TSANN is based on LSTM with the addition of hierarchical attention and elapsed time embeddings.

**Figure 3.**
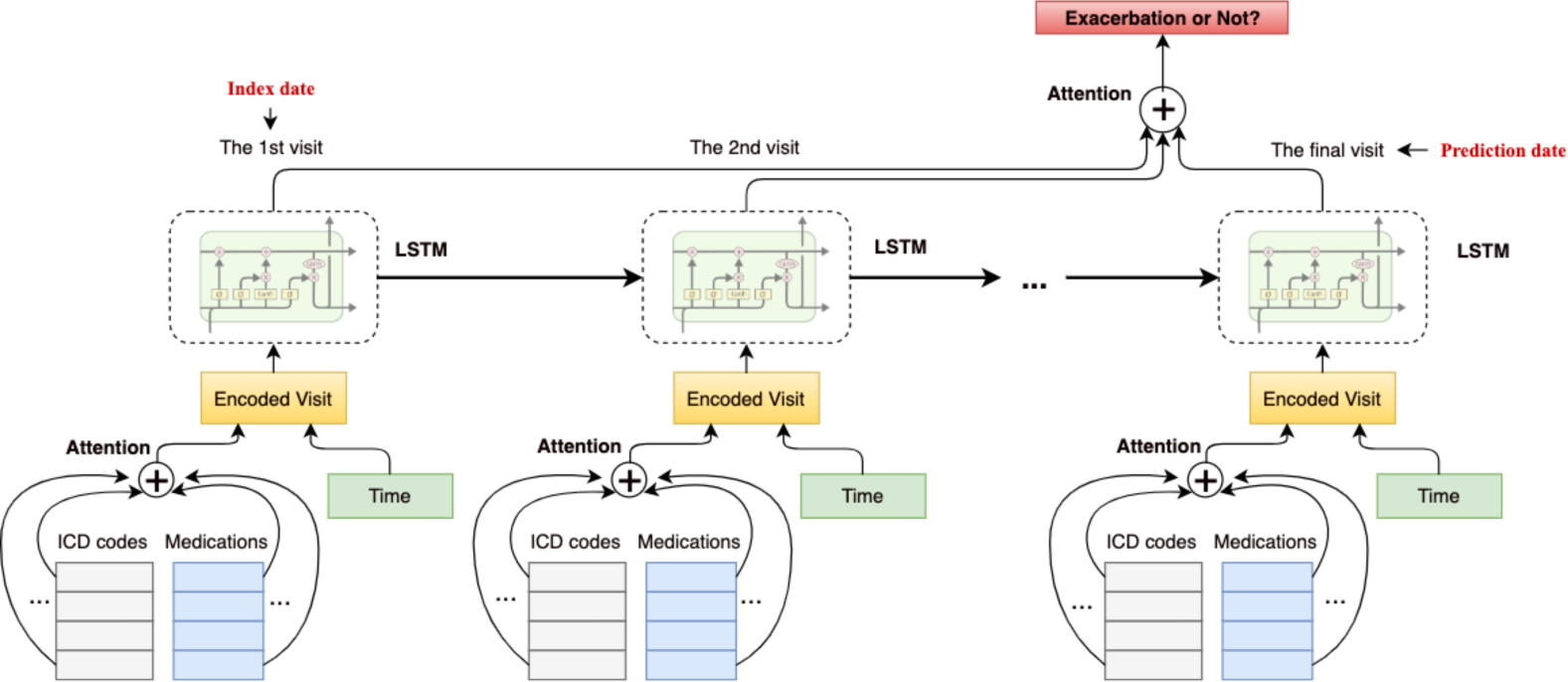
The overview of the TSANN model for asthma exacerbation prediction.

For each visit, multiple clinical variables are encoded in the input layer and averaged through the code-level attention mechanism. The elapsed time embedding is concatenated as the complementary information to indicate the relative time interval between each visit date and the prediction date. LSTM accepts the sequence of encoded visits as inputs and outputs further encodings for each visit. The visit-level attention layer is then applied on the outputs of LSTM to summarize all the visits for each patient. Finally, by feeding the output of visit-level attention into the softmax function, a probability indicating the risk of disease onset is generated.

#### Input

The inputs of the model consist of two types of features. One type is clinical variables including ICD codes, medications, and demographic features. The ICD-10 codes are all converted into ICD-9 based on predefined mappings[31]. All the medications are normalized to their generic names. The demographic features include age, gender, and race, which are only taken as inputs on the visit of the prediction date. Using a projection matrix 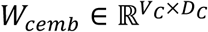, we mapped each clinical variable into a concept embedding vector:

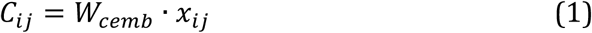

where *C*_*ij*_ is the generated concept embedding vector and 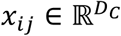 is the one-hot vector denoting the existence of each variable.

The other feature type is time features, which indicate the occurrence time for each clinical variable. Intuitively, variables with different timestamps would behave differently in prediction. For instance, in many cases, a clinical event that happens several days ago would play a more important role than one that happened several months ago. Meanwhile, due to the nature of data irregularity and deficiency of EHRs, successive visits always have diverse time intervals[23], which makes it indispensable to consider the time elapse when doing predictive modeling.

Inspired by the idea of position embeddings in natural language processing which were introduced to model the positional information for each word in e.g. relation classification[32] and the transformer structure in neural language modeling[28,33,34], we introduced elapsed time embeddings to represent the relative time gap for each clinical variable.

Specifically, taking the time of the prediction date *T*_0_ as a pivot, the time feature of each variable is the absolute difference between its occurrence time *T*_i_ and *T*_0_, i.e. *T*_0_ -*T*_i_. Since the observed time window has an upper bound of 365 days, the vocabulary size of the time embeddings was set to be 365. We applied a matrix 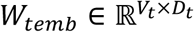 to project each time value to an *m*-dimension vector. Unlike the code embeddings, elapsed time embeddings are fed into the model after the code-level attention and assigned to each visit. The equation to get the elapsed time embedding for each visit is analogous to that for concept embeddings where 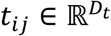

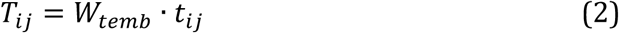

For easier description, we denote each visit as *v*_*i*_, *i* ϵ{1, 2,…,*T*} and each clinical variable in each visit as *v*_*ij*_, *j* ϵ{1, 2,…, *M* }, where *T* is the maximum number of visits and *M* is the maximum number of events in each visit.

#### Code-level Attention

Attention is a mechanism specifically designed for deep neural networks that acts as an information filter, meanwhile has the capacity of alleviating information loss when dealing with long sequences. It selects important sequence spans by assigning weights to different elements in a sequence[35,36]. Through attention, each variable is assigned a weight so that important variables would have larger weights than the others. We adopted the attention mechanism from [37] in which the weight of each variable is generated according to the sequence and a context vector. Concretely, given the set of codes {*v*_*ij*_ }, *j* ϵ{1, 2,…, *M* }in the *i*th visit, the encoded representation of *v*_i_ can be generated by:

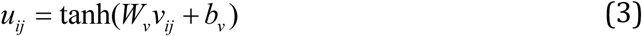

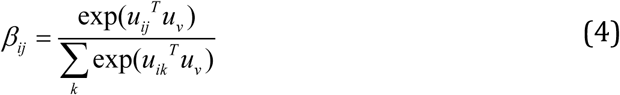

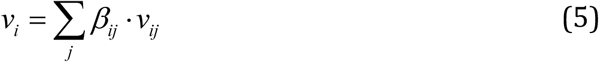

where *W*_v_ and *b*_v_ are the weight and bias for matrix transformation, *u*_ij_ is the attention vector for each code *j* in *v*_i_, *u*_v_ is the context vector for *v*_i_ and is updated during training, and *β*_*ij*_ is the attention weight for the event *v*_*ij*_ based on which we can generate the final weight for this variable.

### Visit-level Attentive LSTM Layer

Taking the encoded representation of each visit as input, LSTM models the sequential information in the observed time window and gets the summarization at the final step (the prediction date). The advantage of LSTMs over basic RNNs is that they can alleviate the vanishing gradient problem, and are thus able to retain “memories” of prior timestamps[38,39]. LSTMs are implemented by several matrix multiplications and nonlinear transformations that aim to mimic the memory mechanism of human brains, in which these operations are called *gates*, signifying that the network can select effective information and abandon useless information. The equations of LSTMs are listed as follows:

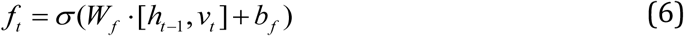

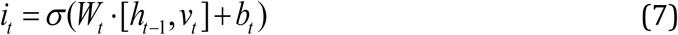

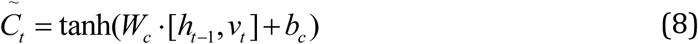

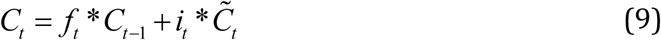

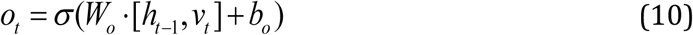

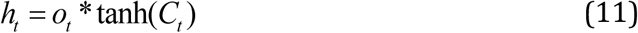

where *W*s and *b*s are weights and biases for different gates or cells (*f*_t_: forget gate, *i*_t_: input gate, *C*_t_: memory cell, *o*_t_:output gate, *h*_t_: hidden cell), and *σ* is the activation function such as *tanh* or *sigmoid*.

We applied self-attention again on the visit-level to measure the risk scores for each clinical variable in each visit. By assigning attention weights to the outputs of LSTM from each step, we can weight each visit in the observed time window.

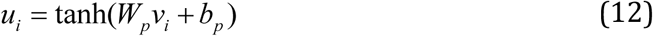

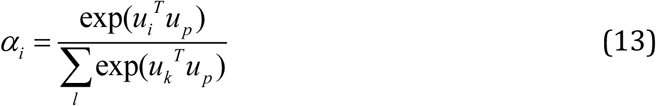

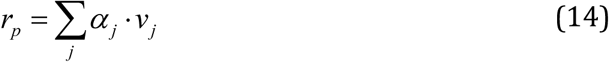

where *W*_p_ and *b*_p_ are the weight and bias for matrix transformation, *u*_i_ is the attention vector for each visit *i* given *v*_i_, *u*_p_ is the context vector, and *a*_j_ is the attention weight for *v*_j_. This process can be seen as a simulation towards the diagnosis procedure of a clinic visit, during which a physician would look back into a patient’s EHR, measure the impacts of each historical clinical event and make a final decision.

### Output

The visit-level attention layer compresses all the information in the observed time window into a fixed size vector. The output of attention further goes through a fully connected layer with a nonlinear activation. A softmax function is finally applied to generate the predicted probability *p*

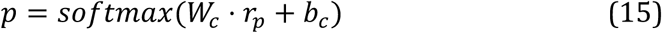

where *r*_*p*_ stands for the output of visit-level attention-LSTM. The value *p* is used as the score for the risk of developing an asthma exacerbation.

### Evaluation

AUROC/AUC (Area Under the Receiver Operating Curve) is widely used as an evaluation metric for predictive models which reflects a balance between sensitivity and specificity[40]. According to the predicted probability *p* (between 0 and 1) for each instance, the AUC value is generated by setting different cut-offs. The methods listed in Table 2 were compared in our experiments.

**Table 2.** The methods used for comparisons.

| Method | Note |
| --- | --- |
| *LR-sparse* | A popular conventional machine learning algorithm[41], usually behaves as a strong baseline in predictive modeling[42]. The input of LR-sparse for each sample is a fixed length feature vector, the length of which is the number of distinct variables (the vocabulary size) and the value of each dimension is the occurrence times of each variable. LR suffered from the data imbalance problem on this dataset, so we employed SMOTE[43] to do over-sampling and help reduce its impact. |
| *LR-dense* | A simplified version of Multi-Layer Perceptron[44] with only one input layer and one softmax layer. In LR-dense, the representations of all the codes were averaged after being projected to the embedding space. The differences between LR-dense and LR-sparse are two-fold: a) the inputs of LR-dense is the average of code embeddings while the input of LR is a fixed length vector denoting the occurrence of each clinical variable; b) the input embeddings of LR-dense can be fine-tuned during training but the input of LR-sparse cannot. |
| *LSTM* | The basic LSTM algorithm, taking the sequence of the clinical variables as input ordered by time; the variables in each visit are averaged. |
| *ALSTM* | Attention LSTM with one layer of LSTM and one layer of attention. |
| *TLSTM*[23] | The time-aware LSTM model, which is one of the state-of-the-art predictive models. In TLSTM, the time gap is used to compute the information decay in the LSTM unit. |
| *RETAIN*[21] | A two-layer attention model, which is another state-of-the-art model for disease onset prediction. In RETAIN, the time features are not embedded as vectors, but real values denoting the gaps from the first visit. |
| *TSANN-I* | The proposed TSANN model but with the second attention layer removed, the prediction is based on the final state of LSTM. |
| *TSANN-I-step* | Use the time encoding method from [34] on TSANN-I, in which although time was also encoded using a vector, it only showed the order of each visit, but not the actual elapsed time, which is insufficient in modeling the irregularity in EHRs. |
| *TSANN-II* | A complete version of the proposed TSANN model. |

For evaluation, we firstly split the data into a training set and a held-out testing set with a ratio of 8:2. Further, 5-fold cross-validation was performed on the training dataset for parameter tuning. During cross-validation, grid search was applied to tune the hyperparameters. Finally, the hyperparameters for our best model TSANN-I were batch size=32, code embeddings dimension=100, time embeddings dimension=20, learning rate=0.001, l2 penalty=0.0001 for all layers, Leaky_ReLU[45] as the activation function for LSTM, adding batch normalization before softmax, and Adam[46] as the optimizer. A more detailed parameter tuning process is shown in the Multimedia Appendix 1. Codes for RETAIN and TLSTM were provided by the respective authors, and all other deep learning models were implemented with TensorFlow[47] and tested on Nvidia Tesla V100, Quadro P6000 and Titan XP GPUs.

## Results

### AUC Values

Since the time information is of critical importance in modeling EHR data, we conducted experiments on situations both with and without it. We did not implement LR-sparse with time embeddings associated with day as the time unit since it would have introduced a greater number of variables (i.e. 12,390*365), which would have been too sparse and difficult for computation. Instead, we reduced the vocabulary of the time variable by setting month as the time unit and finally 148,680 distinct clinical variables were generated. For TLSTM, we only considered a with-time version since it is defined as a time-aware variant of LSTM. For LR-dense, LSTM, ALSTM, TSANN-I and TSANN-II, we used the elapsed time embeddings introduced in this study. The AUC values on the testing set for all the methods are show in Table 3.

**Table 3.** AUC values by the proposed models compared with baselines. (+/-stands for the improvement of adding time info).

| Method | Without time | With time | +/- (%) |
| --- | --- | --- | --- |
| *LR-sparse* | 0.5685 | 0.5825 | +1.4 |
| *LR-dense* | 0.6545 | 0.6753 | +2.08 |
| *LSTM* | 0.6045 | 0.6567 | +5.22 |
| *ALSTM* | 0.6346 | 0.6714 | +3.68 |
| *TLSTM* | - | 0.6548 | - |
| *RETAIN* | 0.6455 | 0.6882 | +4.27 |
| *TSANN-I* | 0.6692 | **0.7003** | +3.11 |
| *TSANN-I-step* | 0.6463 | - | - |
| *TSANN-II* | **0.6827** | 0.6855 | +0.28 |

\*the optimal value for each column is marked in bold.

Compared with different rows, we notice that TSANN-I with time information achieves the optimal AUC value, improving the strongest baseline (RETAIN) by 1.21%. TSANN-II gets comparable performance with RETAIN. All the results show that the conventional machine learning method LR-sparse behaves worse than the deep learning methods. It is noticed that LR-dense performs better on both with and without time than LSTM and ALSTM. TSANN-I-step, which only used time embeddings to denote the relative position of each visit, does not get good result.

When comparing the results with and without time information, considerable improvements were observed after adding time information on most methods, signifying that the problem of data irregularity is obvious in the studied problem and the time information plays an important role in modeling visits for predicting asthma exacerbation. For example, TSANN-I integrated the time interval into decay functions and obtained a 3.11% improvement. Similar cases can also be observed in other models including RETAIN. Surprisingly, TSANN-II, when integrating time embeddings, did not get much improvement. In addition, if without time information, our proposed methods also perform much better than others, showing that they have strengths even in cases such as the loss of temporal information.

Considerable improvements can also be observed on testing set B (RETAIN: 0.7761, TSANN-I: 0.8202) as well as the contribution of adding the time information. And as expected, the general results were much better on testing set B (see Multimedia Appendix 1).

### Personalized Heatmap

In our study, a heatmap conveys the interpretations and behaves as a visualization tool in identifying the personalized risk factors. A heatmap is to illustrate how each candidate risk factor behaves in each visit in the progression of asthma. Each grid in the heatmap is colored based on the attention weights derived from the model. The darker an area is, the more importance the clinical variable signifies, and the more possible it behaves as a risk factor. For example, Figure 4 shows a case where the symptoms of *hypoxemia, shortness of breath* and *wheezing* (799.02, 786.05 and 786.07 in ICD 9), etc. are recognized as possible risk factors. A possible explanation might be the patient’s status of hypoxemia worsened the condition of asthma, following symptoms in breath, and asthma exacerbation was then diagnosed.

**Figure 4.**
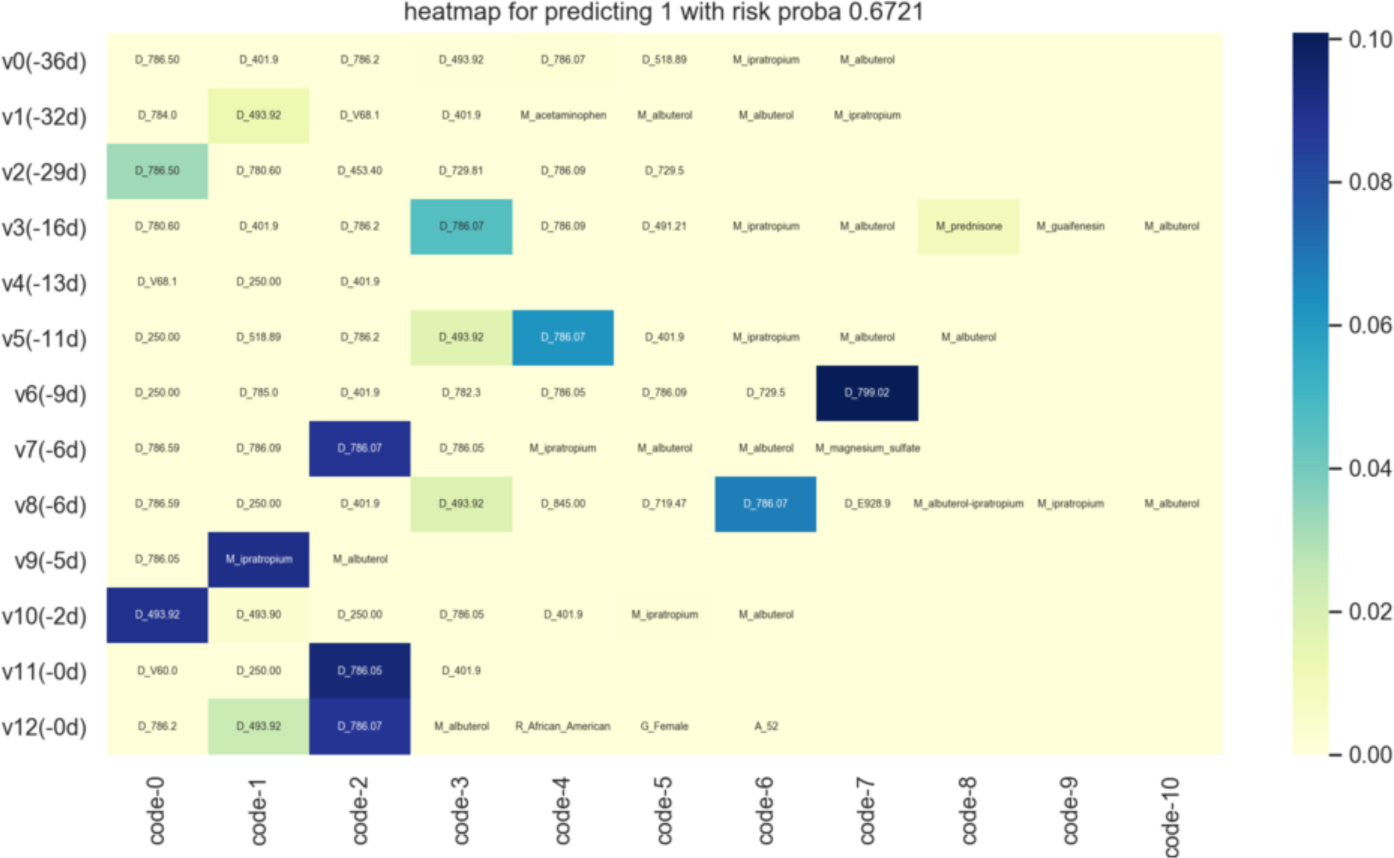
An example of heatmap with the most possible risk factor denoted by the clinical variables: hypoxemia (D_799.02), shortness of breath (D_786.05) and wheezing (D_786.07), etc.

It is hard to get a clear overview of the disease progression since we can only depend on structured data but without any clinical notes. As a result, we can hardly confirm the discovered factors are real risk factors but only know that they might be either possible factors triggering exacerbations or of high associations with the event. However, our method may behave as an important complement in disease prediction and clinical decision.

### Cohort-level Risk Factors

We also proposed a method to discover common risk factors on the population level so that clinicians can have a better understanding towards the disease progression while patients can pay more attention to risk factors in daily lives. The recognized top-ranked risk factors are shown in Table 4. The details of the method and clinical discussion to the factors are described in Multimedia Appendix 1.

**Table 4.**
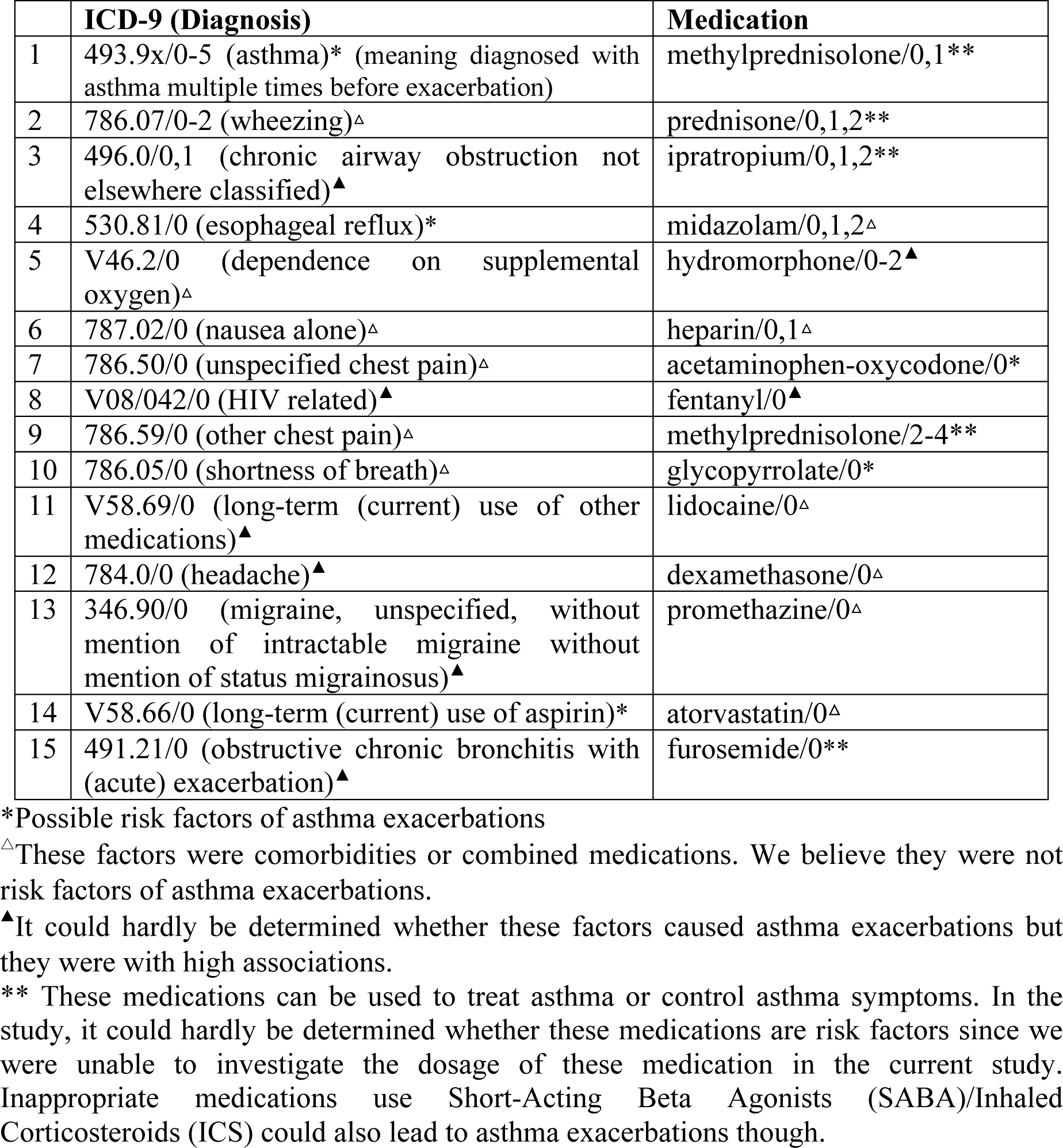
Clinical variables with the top-ranked weights (/N stands for the variable occurred in *N* months prior to the prediction date). We regard both * and ▴ containing valuable information.

|  | ICD-9 (Diagnosis) | Medication |
| --- | --- | --- |
| 1 | 493.9x/0-5 (asthma)* (meaning diagnosed with asthma multiple times before exacerbation) | methylprednisolone/0,1** |
| 2 | 786.07/0-2 (wheezing)<sup>△</sup> | prednisone/0,1,2** |
| 3 | 496.0/0,1 (chronic airway obstruction not elsewhere classified)<sup>▲</sup> | ipratropium/0,1,2** |
| 4 | 530.81/0 (esophageal reflux)* | midazolam/0,1,2<sup>△</sup> |
| 5 | V46.2/0 (dependence on supplemental oxygen)<sup>△</sup> | hydromorphone/0-2<sup>▲</sup> |
| 6 | 787.02/0 (nausea alone)<sup>△</sup> | heparin/0,1<sup>△</sup> |
| 7 | 786.50/0 (unspecified chest pain)<sup>△</sup> | acetaminophen-oxycodone/0* |
| 8 | V08/042/0 (HIV related)<sup>▲</sup> | fentanyl/0<sup>▲</sup> |
| 9 | 786.59/0 (other chest pain)<sup>△</sup> | methylprednisolone/2-4** |
| 10 | 786.05/0 (shortness of breath)<sup>△</sup> | glycopyrrolate/0* |
| 11 | V58.69/0 (long-term (current) use of other medications)<sup>▲</sup> | lidocaine/0<sup>△</sup> |
| 12 | 784.0/0 (headache)<sup>▲</sup> | dexamethasone/0<sup>△</sup> |
| 13 | 346.90/0 (migraine, unspecified, without mention of intractable migraine without mention of status migrainosus)<sup>▲</sup> | promethazine/0<sup>△</sup> |
| 14 | V58.66/0 (long-term (current) use of aspirin)* | atorvastatin/0<sup>△</sup> |
| 15 | 491.21/0 (obstructive chronic bronchitis with (acute) exacerbation)<sup>▲</sup> | furosemide/0** |

\*Possible risk factors of asthma exacerbations
<sup>△</sup>These factors were comorbidities or combined medications. We believe they were not risk factors of asthma exacerbations.
<sup>▲</sup>It could hardly be determined whether these factors caused asthma exacerbations but they were with high associations.
\*\* These medications can be used to treat asthma or control asthma symptoms. In the study, it could hardly be determined whether these medications are risk factors since we were unable to investigate the dosage of these medication in the current study. Inappropriate medications use Short-Acting Beta Agonists (SABA)/Inhaled Corticosteroids (ICS) could also lead to asthma exacerbations though.

## Discussion

### Principle Results

Our proposed method obtains the optimal AUC value on the prediction task, with hierarchical attention and elapsed time embeddings as its booster. The visualization part also provides useful tracks for better understanding the disease progression.

Regarding the AUC values, since we only selected 5 visits as the observed time window, LSTM and ALSTM may not be as powerful as they were in modeling longer sequences, which can be reflected by the unsatisfying results compared with LR-dense. However, for TSANN, where more complex attentive structures were added, the results get comparable with or better than LR-dense. TLSTM, although has also integrated the time decay information, does not get satisfying results, perhaps due to the improper heuristic decaying function for the current dataset. TSANN-I and -II obtain better results as RETAIN, signifying the effectiveness of the model structure. Besides, the hierarchical attention architecture makes it easier for further interpretations, e.g. the personalized heatmap. LR-sparse, although has been tuned thoroughly in our experiment, still behaves worse than the others, which may partly due to its insufficiency in modeling complex sequential patterns.

A typical characteristic of the EHR data is irregularity, which means that clinic visits may be randomly and sparsely distributed along the timeline, and sometimes are even missing. Thus, the predictive model is responsible of serializing the visits for each patient with the consideration of time elapses between continuous visits. The comparisons between results with and without time information in Table 3 demonstrate the effectiveness of considering time elapses on this study cohort. It can be concluded that the prediction of asthma exacerbation is quite time-sensitive, and most of the critical risk factors should have been timestamped. For instance, even for a visit just in front of the prediction date, if the occurrence time of this visit is long before, its impact would still be reduced. Similar case can also be found in [23] with an improvement of 6% from LSTM to TLSTM. For TSANN-I-step, although time embeddings were also used, they were only used to denote the relative position of each visit in the sequence but lack the ability to represent time decay, which can hardly get good results here. Adding time to TSANN-II did not get much improvement as those in other methods, which we think might be attributed to that the addition of the visit-level attention weakens the contribution of the time embeddings.

Apart from the personalized heatmaps and cohort-level risk factors, making use of the weights generated by Eq. 2 and Eq. 3 in Multimedia Appendix 1, we can also visualize how each clinical variable contributes across time, e.g. a variable may behave distinctly among individuals, with different action time or different incidences. Figure 5-6 show two examples in which the time distributions for the clinical variables are displayed through scatters. In these scatters, each circle represents a patient and its size and color depth denote the importance of the corresponding variable for the patient. In the figures, the x-axis stands for the time gap between the occurrence date of the variable to the prediction date, while the y-axis was employed merely for cosmesis. We randomly select a maximum of 2,000 patients to plot this figure.

**Fig 5.**
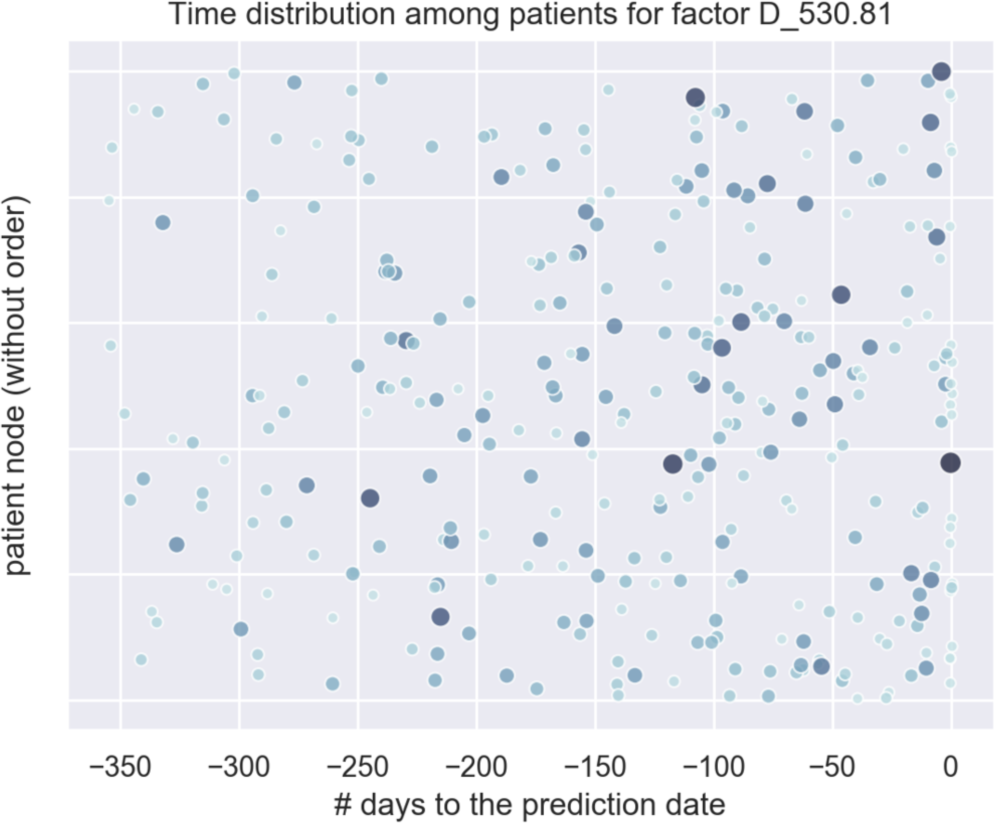
The time distribution of the clinical variable ICD-9:530.81(gastro-esophageal reflux disease) as a risk factor.

**Fig 6.**
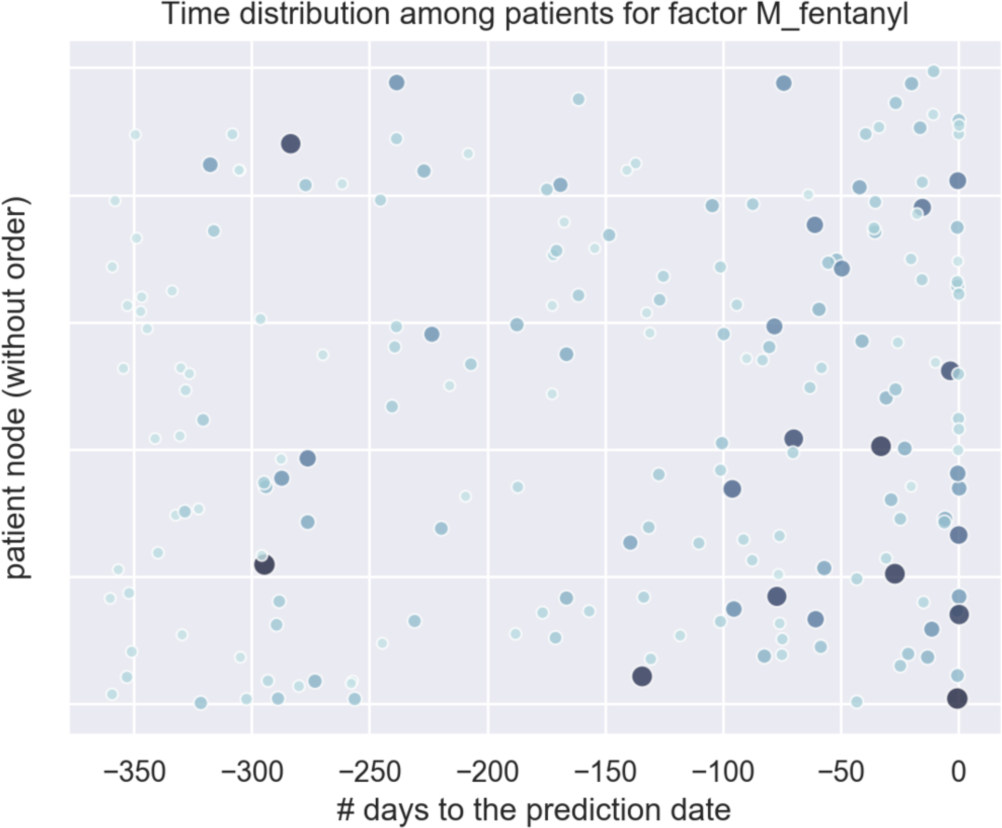
The time distribution of the clinical variable medication: fentanyl as a risk factor.

Figure 5-6 are derived from an ICD code (esophageal reflux: 530.81 in ICD 9) and a medication (fentanyl) respectively. We observe different effective time ranges for these two factors, where the first factor tends to distribute more between the previous 250 to 50 days while the second between the previous 150 days. We hope these visualizations can help figure out the distributions of more possible risk factors to aid the asthma control.

### Comparison with Prior Work

As far as we know, this is the first study in deep learning-based prediction on asthma exacerbation, and from Table 3, we observed that our proposed method outperforms both conventional machine learning and deep learning methods. The performance boosts mainly come from the architecture of hierarchical attentions, which can efficiently capture information from distinct medical elements, and the way of encoding time using elapsed time embeddings, which enables learning temporal patterns from different perspectives. Generally, deep learning-based clinical predictive modeling applied RNN style structures, with minor modifications in structures. For example, [20] used a one-layer RNN and [21] used a two-layer RNN with their attention weights but one attention is for evaluating each embedding dimension. In comparison, we applied attention weights on both the code and visit level, which is also easy for the interpretation of results.

Strategies of encoding time in previous studies can be generally categorized into learning a subspace decomposition of the cell memory in RNNs to enable time decay[20,23] or taking the time values as features[21,48]. These methods, since only used time as a single value, limited the representation ability of time if multiple possible patterns exist in the timeline, e.g. a clinical event *c* happened in time *t* might be modeled jointly in causal-like patterns such as *a*_t1_->*c*_t_ and *c*_t_->*b*_t2_, in which *t* may behave differently.

### Limitations and Future Work

By using deep learning, we offered a novel way of identifying possible risk factors and predicting the risk of asthma exacerbation. However, the current work still has some limitations. *First*, for the model interpretation part, how multiple clinical variables interact with each other needs further exploration, simply considering each variable independently may loss the dependency patterns between them, e.g. the prescription of a drug might be closely associated with a disease or symptom. *Secondly*, EHRs have their own drawbacks such as data irregularity, sparsity, and noise. Some potential risk factors of asthma exacerbations might not be recorded in EHRs. As a result, the information integrity cannot be well guaranteed. We may need to find ways to make the data more complete such as including information from textual reports or patient surveys. *Finally*, the performance of the model still has room to improve. It might be boosted further by designing more powerful structures or including background knowledge.

## Conclusion

In this paper, we proposed an attentive deep learning network for asthma exacerbation prediction and employed elapsed time embeddings to model the time decays. By leveraging the weights of the model, we not only generated personalized heatmaps and specific risk scores at the individual-level, but also identified possible risk factors of asthma exacerbation at the cohort-level. Compared with previous studies, our model is effective in modeling time information and obtains better overall AUCs. Since the model is completely data-driven and relies little on feature engineering, it can easily be generalized to other prediction tasks. To our best knowledge, this is the first study to predict asthma exacerbation risks using a deep learning model and includes elaspsed time embeddings. Some of the top-ranked risk factors identified have gained supporting evidence from previous medical researches, which proved our method has good reliability and accuracy.

## Data Availability

The Cerner data used is available for external users if sponsored by UTHealth SBMI faculty, have a UTHealth account, and have signed a Cerner data user agreement that is co-signed by the sponsoring SBMI faculty member.

https://sbmi.uth.edu/sbmi-data-service/data-set/cerner/

## Acknowledgements

CT conceived the research project. YX, HJ and CT designed the pipeline and method. YX implemented the deep learning model of the study and prepared the manuscript. HJ completed the clinical part of the manuscript. WJZ and HX provided valuable suggestions on the cohort selection and experiment design. YZhou and YZhang extracted, cleaned the data and did statistics. LR helped reorganize the data and did normalizations for the revised version. FL, JD, SW, DZ and CT proofread the paper and provided valuable suggestions. All the authors have read and approved the final manuscript.

We thank Dr. Irmgard Willcockson for proofreading. Also, thanks to Cerner for providing the valuable Health Facts EMR data. We gratefully acknowledge the support of NVIDIA Corporation with the donation of the Quadro P6000 and TITAN XP GPUs used for this research.

This research was partially supported by the National Library of Medicine of the National Institutes of Health under award number R01LM011829, the National Institute of Allergy and Infectious Diseases of the National Institutes of Health under award number 1R01AI130460, National Center for Advancing Translational Sciences of the National Institutes of Health under award number U01TR02062, and the Cancer Prevention Research Institute of Texas (CPRIT) Training Grant #RP160015.

## Abbreviations

ALSTM: Attention Long Short-Term Memory
AUROC/AUC: Area Under the Receiver Operating Curve
EHRs: Electronic Health Records
ICS: Inhaled Corticosteroids
LR: Logistic Regression
LSTM: Long Short-Term Memory
RETAIN: REverse Time AttentIoN model
RNNs: Recurrent Neural Networks
SABA: Short-Acting Beta Agonists
SMOTE: Synthetic Minority Over-sampling Technique
TLSTM: Time-aware Long Short-Term Memory
TSANN: Time-Sensitive Attentive Neural Network

## References

1 Organization WH. Asthma. http://www.who.int/mediacentre/factsheets/fs307/en/.

2 CDC. Most Recent Asthma Data. https://www.cdc.gov/asthma/most_recent_data.htm.

3 GINA. Pocket Guide for Asthma Management. Pocket Guid asthma Manag Prev 2018.

4 Nurmagambetov T, Kuwahara R, Garbe P. The economic burden of asthma in the United States, 2008-2013. Ann Am Thorac Soc 2018;15:348–56. doi: 10.1513/AnnalsATS.201703-259OC

5 Wark PAB, Gibson PG. Asthma exacerbations · 3: Pathogenesis. Thorax 2006;61:909–15. doi: 10.1136/thx.2005.045187

6 Levy ML. The national review of asthma deaths: What did we learn and what needs to change? Breathe 2015;11:15–24. doi: 10.1183/20734735.008914

7 Fleming L. Asthma exacerbation prediction. Curr Opin Allergy Clin Immunol 2018;:1. doi: 10.1097/ACI.0000000000000428

8 Azizpour Y, Delpisheh A, Montazeri Z, et al. Effect of childhood BMI on asthma: A systematic review and meta-analysis of case-control studies. BMC Pediatr 2018;18:1–13. doi: 10.1186/s12887-018-1093-z

9 Lieu TA, Quesenberry CP, Sorel ME, et al. Computer-based models to identify high-risk children with asthma. Am J Respir Crit Care Med 1998;157:1173–80. doi: 10.1164/ajrccm.157.4.9708124

10 Stanford RH, Nagar S, Lin X, et al. Use of ICS/LABA on Asthma Exacerbation Risk in Patients Within a Medical Group. J Manag care Spec Pharm 2015;21:1014–9. doi: 10.18553/jmcp.2015.21.11.1014

11 Bzdok D, Altman N, Krzywinski M. Statistics versus machine learning. Nat Publ Gr 2018;15:233–4. doi: 10.1038/nmeth.4642

12 Dexheimer JW, Brown LE, Leegon J, et al. Comparing Decision Support Methodologies for Identifying Asthma Exacerbations. 2007;:880–4.

13 Jeong JF, Cheol I. Machine learning approaches to personalize early prediction of asthma exacerbations. Ann N Y Acad Sci 2017;1387:153–65. doi: 10.1016/j.coviro.2015.09.001.Human

14 Sanders DL, Aronsky D. Detecting Asthma Exacerbations in a Pediatric Emergency Department Using a Bayesian Network. 2006;:684–8.

15 Loymans RJB, Debray TPA, Honkoop PJ, et al. Exacerbations in Adults with Asthma: A Systematic Review and External Validation of Prediction Models. J Allergy Clin Immunol Pract 2018;6:1942-1952.e15. doi: 10.1016/j.jaip.2018.02.004

16 Bae SH, Choi I, Kim NS. Acoustic Scene Classification Using Parallel Combination of LSTM and CNN. Proc Detect Classif Acoust Scenes Events 2016 Work 2016;:11–5.

17 Lecun Y, Bengio Y, Hinton G. Deep learning. Nature 2015;521:436–44. doi: 10.1038/nature14539

18 Baytas IM, Xiao C, Zhang X, et al. Patient Subtyping via Time-Aware LSTM Networks. Proc 23rd ACM SIGKDD Int Conf Knowl Discov Data Min - KDD ‘17 2017;:65–74. doi: 10.1145/3097983.3097997

19 Xiao C, Choi E, Sun J. Opportunities and challenges in developing deep learning models using electronic health records data: a systematic review. J Am Med Informatics Assoc 2018;00:1–10. doi: 10.1093/jamia/ocy068

20 Rajkomar A, Oren E, Chen K, et al. Scalable and accurate deep learning for electronic health records. Published Online First: 2018. doi: 10.1038/s41746-018-0029-1

21 Choi E, Bahadori MT, Kulas JA, et al. RETAIN: An Interpretable Predictive Model for Healthcare using Reverse Time Attention Mechanism. Published Online First: 2016.http://arxiv.org/abs/1608.05745

22 Ma F. Dipole : Diagnosis Prediction in Healthcare via Attention-based Bidirectional Recurrent Neural Networks. 2017.

23 Baytas IM, Xiao C, Zhang X, et al. Patient Subtyping via Time-Aware LSTM Networks. Proc 23rd ACM SIGKDD Int Conf Knowl Discov Data Min - KDD ‘17 2017;:65–74. doi: 10.1145/3097983.3097997

24 Wu S, Liu S, Sohn S, et al. Modeling Asynchronous Event Sequences with RNNs. J Biomed Inform Published Online First: 2018. doi: 10.1016/j.jbi.2018.05.016

25 Che C. An RNN Architecture with Dynamic Temporal Matching for Personalized Predictions of Parkinson ‘ s Disease.

26 Jin H, Young H. Learning representations for the early detection of sepsis with deep neural networks. Comput Biol Med 2017;89:248–55. doi: 10.1016/j.compbiomed.2017.08.015

27 Jin H, Young H. Learning representations for the early detection of sepsis with deep neural networks. Comput Biol Med 2017;89:248–55. doi: 10.1016/j.compbiomed.2017.08.015

28 Vaswani A, Shazeer N, Parmar N, et al. Attention Is All You Need. Published Online First: 2017. doi: 10.1017/S0952523813000308

29 GINA. Global Strategy For Asthma Management and Prevention. Glob Initiat Asthma 2017;:http://ginasthma.org/2017-gina-report-global-strat. xdoi: 10.1183/09031936.00138707

30 Bai TR, Vonk JM, Postma DS, et al. Severe exacerbations predict excess lung function decline in asthma. Eur Respir J 2007;30:452–6. doi: 10.1183/09031936.00165106

31 Mapping between ICD-10 and ICD-9. https://www.health.govt.nz/nz-health-statistics/data-references/mapping-tools/mapping-between-icd-10-and-icd-9 (accessed 1 Jan 2019).

32 Daojian Zeng, Kang Liu, Siwei Lai GZ and JZ. Relation Classification via Convolutional Deep Neural Network. In: Proceedings of COLING 2014, the 25th International Conference on Computational Linguistics: Technical Papers. 2014. 2335–44. doi: 10.1021/bi990527s

33 Alec R, Karthik N, Tim S, et al. Improving Language Understanding by Generative Pre-Training. OpenAI 2018;:1–10. doi: 10.1093/aob/mcp031

34 Song H, Rajan D, Thiagarajan JJ, et al. Attend and diagnose: Clinical time series analysis using attention models. 32nd AAAI Conf Artif Intell AAAI 2018 2018;:4091–8.

35 Bahdanau D, Cho K, Bengio Y. Neural Machine Translation by Jointly Learning to Align and Translate. 2014;:1–15. doi: 10.1146/annurev.neuro.26.041002.131047

36 Xiang Y, Chen Q, Wang X, et al. Answer Selection in Community Question Answering via Attentive Neural Networks. IEEE Signal Process Lett 2017;24:505–9. doi: 10.1109/LSP.2017.2673123

37 Yang Z, Yang D, Dyer C, et al. Hierarchical Attention Networks for Document Classification. Proc 2016 Conf North Am Chapter Assoc Comput Linguist Hum Lang Technol 2016;:1480–9. doi: 10.18653/v1/N16-1174

38 Hochreiter S, Urgen Schmidhuber J. Long Short-Term Memory. Neural Comput 1997;9:1735–80. doi: 10.1162/neco.1997.9.8.1735

39 Sak H, Senior A, Beaufays F. Long short-term memory recurrent neural network architectures for large scale acoustic modeling. Interspeech 2014 2014;:338–42. doi: 1402.1128

40 Mandic S, Go C, Aggarwal I, et al. Relationship of predictive modeling to receiver operating characteristics. J Cardiopulm Rehabil Prev 2008;28:415–9. doi: 10.1097/HCR.0b013e31818c3c78

41 Hosmer Jr, David W.,Stanley Lemeshow and RXS. Applied logistic regression. 2013.

42 Choi E, Bahadori MT, Kulas JA, et al. RETAIN: an interpretable predictive model for healthcare using reverse time attention mechanism. Published Online First: 2016.http://arxiv.org/abs/1608.05745

43 Chawla, Nitesh V and Bowyer, Kevin W and Hall, Lawrence O and Kegelmeyer WP. SMOTE: Synthetic Minority Over-sampling Technique Nitesh. J Artif Intell Res 2002;16:321–57. doi: 10.1613/jair.953

44 Pal, Sankar K and Mitra S. Multilayer Perceptron, Fuzzy Sets, Classifiaction. IEEE Trans Neural Networks 1992;3:683 696.

45 Xu B, Wang N, Chen T. Empirical Evaluation of Rectified Activations in Convolution Network. 2015.

46 Kingma DP, Ba J. Adam: a method for stochastic optimization. In: Iclr. 2015. doi:http://doi.acm.org.ezproxy.lib.ucf.edu/10.1145/1830483.1830503

47 Martín Abadi, Ashish Agarwal PB et al. TensorFlow: Large-Scale Machine Learning on Heterogeneous Distributed Systems. 2015. doi: 10.1093/library/s4-X.3.339

48 Rajkomar A, Oren E, Chen K, et al. Scalable and accurate deep learning for electronic health records. npj Digit Med 2018;:1–10. doi: 10.1038/s41746-018-0029-1

